# Modelling COVID-19 evolution in Italy with an augmented SIRD model using open data

**DOI:** 10.1101/2021.04.14.21255500

**Authors:** Vincenzo Nardelli, Giuseppe Arbia, Andrea Palladino, Luigi Giuseppe Atzeni

## Abstract

We propose an augmented version of the traditional SIRD epidemic model and we estimate its parameters using the SaRs-Cov-2 Italian open-data. The model’s parameters are estimated partly using numerical optimization and partly with ABC. Our estimation procedure provides a good fit to real data.

## 1 Introduction

The recent SARS-COV-2 epidemic is the first global pandemic in the big data era. Differently from other past epidemics, it developed even in technologically advanced countries and put the most innovative health systems in crisis. Moreover, this event brought to light different problems related to the quality of data and the related decision-making. Indeed, the public sector in most countries was not ready to collect, validate and distribute open data(Hua and Shaw, 2020) and the lack of statistical knowledge in the citizens and in most of the media led to the inability to clearly distinguish between “data” and “information” (Arbia and Nardelli, 2020) (Zarocostas, 2020). A large number of researchers during the Covid pandemic have unsuccessfully required the access to anonymous individual data. Many active research groups, (among which e. g. Covstat (2020)) developed models to predict the trend of the epidemic using all available open data, trying to mitigate the problems due to the poor data quality and to implement and estimate the model’s parameters together with its uncertainty. In the next section we will present our proposal.

## 2 Model definition

Historically, one of the first model used to predict the spread of the pandemic was the SIR model (Kermack and McKendrick, 1991) based on a system of ordinary differential equations that models 3 categories of population (Susceptible, Infected, Recovered). In any given moment of time *t, I*(*t*) and *S*(*t*) indicate respectively the number of infected people and the number of vulnerable people, while *R*(*t*) (removed) represents the total of those who develop immunity (recovered) or died. Obviously in any moment of time we have: *t, S*(*t*) + *I*(*t*) + *R*(*t*) = *N* with *N* the total population. The SIR model describes the variation of *S*(*t*), *I*(*t*), and *R*(*t*) and the transitions from one category to the other. The original model specification does not consider population mobility in response to possible lockdown measures nor the impact of the asymptomatic. In this paper we propose an adaptation of Khailaie et al. (2020) model which can be applied to model the spread of the epidemic in Italy using the available open-data diffused from Protezione Civile (Palladino et al., 2020).

Our model is based on 6 categories, namely: **S**usceptible (people that can still be affected by the virus): **I**nfected (people that are currently infected); **H**ospitalized (people that need a medical treatment in hospital);I**C**U (people with severe symptoms that need to go to Intensive Care);**R**ecovered (people that recovered from the illness) and **D**eaths. We will refer to this model with the acronym “SIHCRD”. whose schematic representation is reported in Fig.1.

**Fig. 1.**
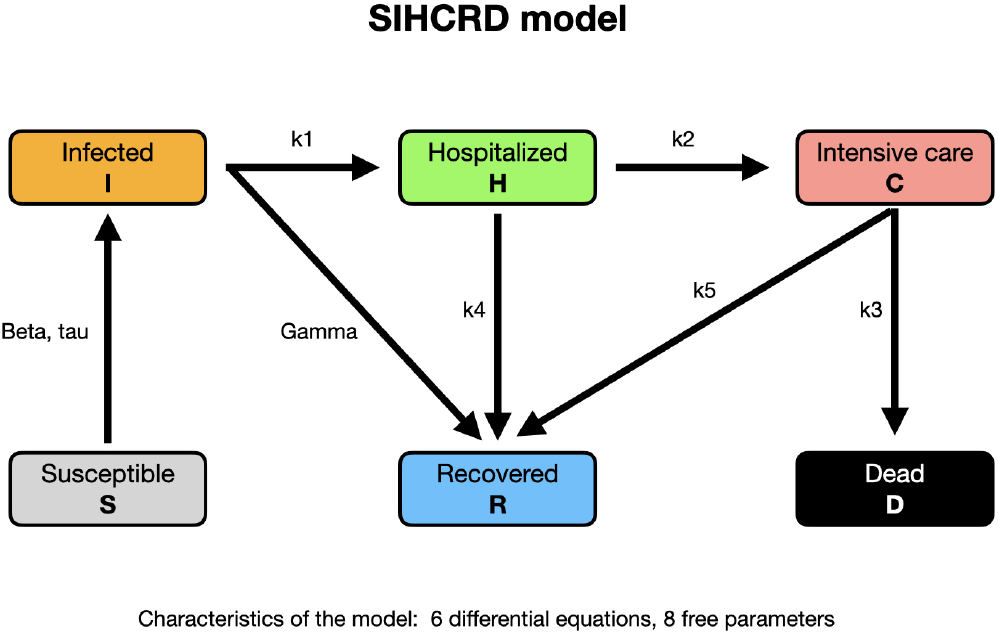
Schematic representation of the model

The model is characterized by six non-linear ordinary differential equations:

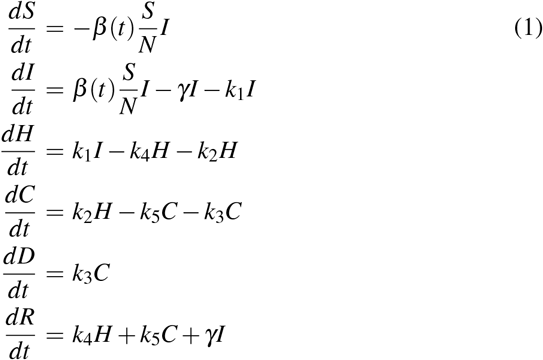

where *β* (*t*) = *β*_0_ · *e*^−*t/τ*^. The model is characterized by 8 free parameters. From the previous equations, we have *S* + *I* + *H* + *C* + *D* + *R* |= *constant*. In what follows we describe the parameters in details.

- *β* is related to the spread of the infection. Larger values of *β* corresponds to a fast spread of the epidemic;
- *γ* is related to the (inverse of) time necessary to move from the category “infected”to the category “recovered”, without passing through hospital;
- *k*_1_ is the product between the fraction of infected people that need to go to hospital (roughly 5% in the Italian experience) times the inverse of the average time required to move from “infected”to “hospitalized”;
- *k*_2_ denotes the product between the fraction of hospitalized people that need to go to intensive care units (roughly 10% in Italy) times the inverse of the average time required to move from “hospitalized”to “intensive care units”;
- *k*_3_ denotes the product between the fraction of patients that die times the average time that they stay in ICU (Intensive Care Unit) before the death;
- *k*_4_ denotes the product between the fraction of people that do not go to ICUs (roughly 90%) times the inverse of the average time required to recover;
- *k*_5_ denotes the product between the fraction of people the do not die in ICUs (roughly 70% during the second pandemic wave in Italy) times the average time required to recover;
- the parameter *τ* denotes the timescale of the decreasing of the parameter *β*

When *k*_1_ = 0 we go back to the original SIR model. The model contains some working hypotheses. The first is that people die only in ICUs. The second is that once a patients is recovered is removed from the susceptible, it it cannot be infected again (Wajnberg et al., 2020). As in the original SIR model the basic reproduction number, usually called *R*_0_, can be obtained combining some of the previous parameters as follows:

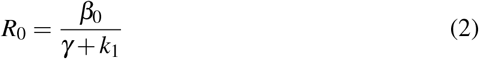

The evolution of *R*_0_ through time, usually indicated as *R*_*t*_, is instead given by:

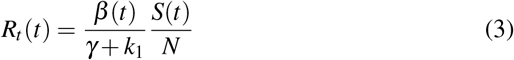

The basic reproduction number gives important indications about the behavior of the epidemic. Indeed, when *R*_*t*_ *>* 1 the epidemic is expanding, when *R*_*t*_ = 1 the epidemic has reached the maximum, while for *R*_*t*_ *<* 1 the epidemic is in the decreasing phase.

## 3 Fitting procedure

In our study we fitted model (1) to the real Italian data during the second wave of the epidemic in the period October 1st and November 15th 2020. For the initial number of infected people we assumed the 6 million, estimated by Bassi et al. (2020). All the others variables in Equation (1) are initialized according to the data available for the previous day. In the loss function, we assign the same weights of the errors (absolute percentage error) to hospitalized (H), patients in ICU (C) and deaths (D), while we don’t use the number of infected people. This procedure allows us to make a more reliable estimate given the large uncertainties in evaluating the number of positive individuals and the irregularities in the transmission of data and in the testing procedures. A mixed approach was used to estimate the model parameters. For the parameters involving the transition between the categories of infected, the model was fitted through numerical optimization starting from the estimates published by the Istituto Superiore di Sanità (ISS) ^1^ and other studies such as Richardson et al. (2020) and Grasselli et al. (2020). In particular, we used the optimizer algorithm Nelder-Mead (Gao and Han, 2012) implemented in SciPy (Virtanen et al., 2020) to tune 6 parameters of the model. In Table 1, we report the result of this optimization.

**Table 1.** Parameter fit after numerical optimization

| Parameter | Fitted value |
| --- | --- |
| $\gamma$ | 0.200 |
| $k_1$ | 0.008 |
| $k_2$ | 0.030 |
| $k_3$ | 0.172 |
| $k_4$ | 0.119 |
| $k_5$ | 0.118 |

The remaining parameters (*β* and *τ*) were estimated through the Approximate Bayesian Computation - Sequential Monte Carlo (ABCSMC) (Toni and Stumpf, 2010) which allows to evaluate the uncertainty of the parameters considering the SIHCRD model as a black-box, starting from non-informative prior. In particular, the prior distribution was assumed to be Uniform between 0 and 1 for the *β* parameter and uniform varies between 0 and 600 for the *τ* parameter. Fig. 2, shows the credible intervals of the estimations using PyABC (Klinger et al., 2018) with a population size of 400 and the stopping rule with minimum error set to 1.5%.In Table 2, we report the estimation of the parameters from the posterior distribution.

**Fig. 2.**
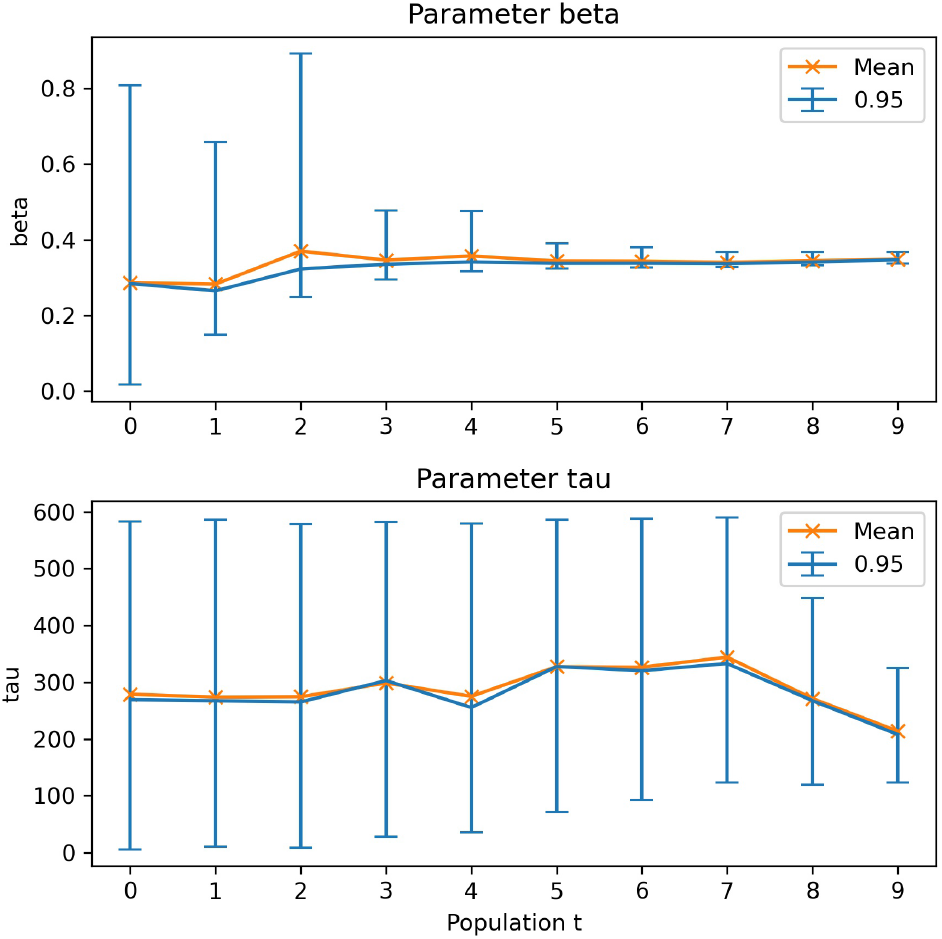
Estimates and credible interval of the parameters *β* and *τ*

**Table 2.**
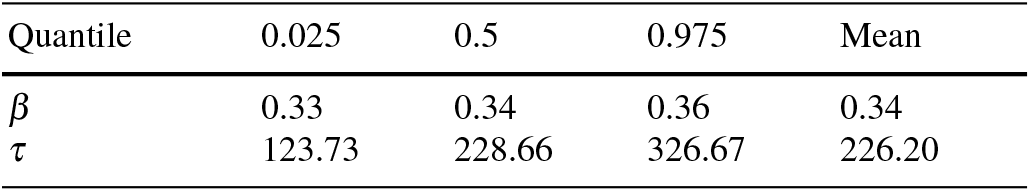
Summary of the posterior distributions

## 4 Estimation results

Details about the proposed model are published online^2^ where the results are constantly updated. During the second wave, the average error of the fit never exceeded 10% despite the great irregularity in the official data. The results are in agreement with other models published during the second wave.

The parameters’ estimation with ABCSMC allows to model the uncertainty. An example of the fit of the hospitalized curve (H) iss reported in Figure 3.

**Fig. 3.**
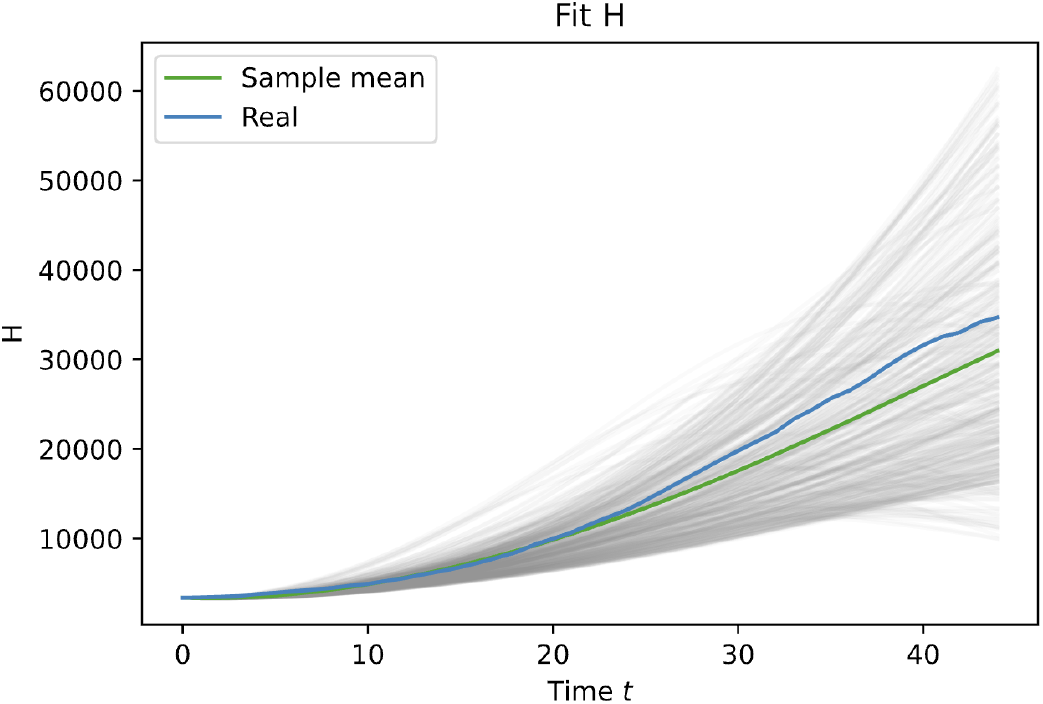
True and estimated of the posterior distribution of hospitalized patients (H)

## 5 Conclusions

In this paper we propose a mixed strategy to estimate the parameters of an augmented SIRD model combining numerical optimization and ABC procedures. In this way we can calculate credible intervals for the crucial epidemic parameters thus helping their interpretation and their use in the monitoring and surveillance of the pandemic diffusion.

## Data Availability

Open data

## Footnotes

1 https://www.epicentro.iss.it/coronavirus/sars-cov-2-decessi-italia

2 https://dashboard.covstat.it/

## References

Giuseppe Arbia and Vincenzo Nardelli. I dati non parlano da soli: l’epoca del Coronavirus smaschera l’inganno dell’algoritmo-onnipotente e rivaluta il metodo statistico. In Giust. Insieme, number 923. 2020. ISBN 978-88-548-2217-7.

F Bassi, G Arbia, and PD Falorsi. Observed and estimated prevalence of covid-19 in italy: How to estimate the total cases from medical swabs data. Science of The Total Environment, page 142799, 2020.

Covstat. Covstat - monitoraggio covid-19 in italia, Nov 2020. URL https://covstat.it/.

Fuchang Gao and Lixing Han. Implementing the nelder-mead simplex algorithm with adaptive parameters. Computational Optimization and Applications, 51(1): 259–277, 2012.

Giacomo Grasselli, Alberto Zangrillo, Alberto Zanella, Massimo Antonelli, Luca Cabrini, Antonio Castelli, Danilo Cereda, Antonio Coluccello, Giuseppe Foti, Roberto Fumagalli, et al. Baseline characteristics and outcomes of 1591 patients infected with sars-cov-2 admitted to icus of the lombardy region, italy. Jama, 323 (16):1574–1581, 2020.

Jinling Hua and Rajib Shaw. Corona virus (covid-19)”infodemic” and emerging issues through a data lens: The case of china. International journal of environmental research and public health, 17(7):2309, 2020.

W. O. Kermack and A. G. McKendrick. Contributions to the mathematical theory of epidemics-I. Bull. Math. Biol., 53(1-2):33–55, mar 1991. ISSN 00928240. doi: 10.1007/BF02464423. URL https://link.springer.com/article/10.1007/BF02464423.

Sahamoddin Khailaie, Tanmay Mitra, Arnab Bandyopadhyay, Marta Schips, Pietro Mascheroni, Patrizio Vanella, Berit Lange, Sebastian Binder, and Michael Meyer-Hermann. Development of the reproduction number from coronavirus sars-cov-2 case data in germany and implications for political measures. medRxiv, 2020.

Emmanuel Klinger, Dennis Rickert, and Jan Hasenauer. pyabc: distributed, likelihood-free inference. Bioinformatics, 34(20):3591–3593, 2018.

Andrea Palladino, Vincenzo Nardelli, Luigi Giuseppe Atzeni, Nane Cantatore, Maddalena Cataldo, Fabrizio Croccolo, Nicolas Estrada, and Antonio Tombolini. Modelling the spread of covid19 in italy using a revised version of the sir model. arXiv preprint 2005.08724, 2020.

Safiya Richardson, Jamie S Hirsch, Mangala Narasimhan, James M Crawford, Thomas McGinn, Karina W Davidson, Douglas P Barnaby, Lance B Becker, John D Chelico, Stuart L Cohen, et al. Presenting characteristics, comorbidities, and outcomes among 5700 patients hospitalized with covid-19 in the new york city area. Jama, 323(20):2052–2059, 2020.

Tina Toni and Michael PH Stumpf. Simulation-based model selection for dynamical systems in systems and population biology. Bioinformatics, 26(1):104–110, 2010.

Pauli Virtanen, Ralf Gommers, Travis E. Oliphant, Matt Haberland, Tyler Reddy, David Cournapeau, Evgeni Burovski, Pearu Peterson, Warren Weckesser, Jonathan Bright, Stéfan J. van der Walt, Matthew Brett, Joshua Wilson, K. Jarrod Millman, Nikolay Mayorov, Andrew R. J. Nelson, Eric Jones, Robert Kern, Eric Larson, CJ Carey, İlhan Polat, Yu Feng, Eric W. Moore, Jake VanderPlas, Denis Laxalde, Josef Perktold, Robert Cimrman, Ian Henriksen, E. A. Quintero, Charles R. Harris, Anne M. Archibald, António H. Ribeiro, Fabian Pedregosa, Paul van Mulbregt, and SciPy 1.0 Contributors. SciPy 1.0: Fundamental Algorithms for Scientific Computing in Python. Nature Methods, 17:261–272, 2020. doi: 10.1038/s41592-019-0686-2.

Ania Wajnberg, Fatima Amanat, Adolfo Firpo, Deena R Altman, Mark J Bailey, Mayce Mansour, Meagan McMahon, Philip Meade, Damodara Rao Mendu, Kimberly Muellers, et al. Robust neutralizing antibodies to sars-cov-2 infection persist for months. Science, 370(6521):1227–1230, 2020.

John Zarocostas. How to fight an infodemic. The Lancet, 395(10225):676, 2020.

